# Variability of Changes in Emotion Recognition with Deep Brain Stimulation Depends on the Location of the Stimulation Volume in the Subthalamic Nucleus

**DOI:** 10.1101/2025.11.03.25339378

**Authors:** Clara Klimaschewski, Dilara Bingöl, Josefine Waldthaler, David Pedrosa

**Affiliations:** Department of Neurology, University Hospital of Marburg and Gießen, Marburg, Germany; Centre of Mind, Brain and Behaviour, Philipps-University Marburg, Marburg, Germany; Department of Clinical Neuroscience, Karolinska Institutet, Stockholm, Sweden

**Keywords:** Parkinson’s disease, STN-DBS, subthalamic nucleus, facial emotion recognition, non-motor symptoms, Volume of activated tissue

## Abstract

The impact of Deep Brain Stimulation (DBS) on emotion processing remains debated. The aim of this study was to investigate whether the localisation of stimulation within the subthalamic nucleus (STN) differentially influences emotion processing in sixteen patients with Parkinson’s disease. An emotion recognition task was employed before and three months after STN-DBS surgery in off-medication state. Volumes of activated tissue (VATs) were mapped to STN subregions and correlated with task performance changes. Larger right ventrolateral non-motor STN VATs were associated with improved recognition of positive emotions (ρ = 0.54, p = 0.04). This association was right-lateralised, involving fibres connecting the STN to the prefrontal cortex, suggesting a lateralised network basis for emotion processing in PD. Larger VATs in left dorsolateral STN were linked to perceiving emotions more positively. Our findings highlight the role of stimulation site in non-motor STN-DBS outcomes which may explain previous inconsistent findings regarding DBS-effects on emotion processing.

## 1. Introduction

Beyond the hallmark motor symptoms of Parkinson’s disease (PD), many patients contend with disabling non-motor symptoms, including affective disorders, REM sleep disturbances, and cognitive impairment [1,2]. Among these, deficits in emotion recognition and their association with social difficulties are increasingly recognised as clinically relevant [3]. These impairments also highlight the complex nature of PD, necessitating a multifaceted management approach. While pharmacological therapies relieve motor symptoms, their effectiveness diminishes over time [4,5]. In these cases, Deep Brain Stimulation (DBS) has emerged as a highly efficacious therapeutic option.

Irrespective of the marked improvements in motor function after surgery [6–8], DBS can both improve and deteriorate non-motor symptoms. There have been reports of enhanced quality of life and improved anxiety, sleep, and pain alongside studies reporting hypomania, apathy, increased impulsivity, and even suicidal ideation after surgery [9–12]. Notably, several studies suggest that DBS especially in the subthalamic nucleus (STN) may disrupt facial emotion recognition (FER), particularly for negative emotions such as fear, sadness, and anger [3,12]. Despite these findings, the evidence for a generalised impairment in FER remains inconclusive, as other studies, including a recent large-scale trial, report no significant impact of DBS on FER one year after lead implantation [13]. Given that emotion recognition is fundamental to human social interaction, particularly in enabling non-verbal communication, fostering empathy, and enhancing interpersonal connections, understanding the impact of DBS on FER is of great importance [3,14,15].

Inconsistencies in what effects are caused by subthalamic DBS on FER, may be attributed to different factors modulating this relationship. First, patient specific variables, such as medication state [8], symptom severity, affective symptoms, executive function, and visuospatial abilities, have to be contemplated [12]. Secondly, methodological variations - including differences in stimulus modality, the applied paradigms, and the specific emotional expressions assessed - may also account for the observed heterogeneity [12,15]. Based on the growing evidence for a role of the STN in emotion processing [16], one might posit that the precise anatomical location of active DBS contacts within the STN creating the volume of activated tissue (VAT) [8] contributes to the variability, as well. This aligns with the known functional compartmentalisation of the STN into sensorimotor, limbic, and cognitive-associative subregions [17,18].

While prior research has established that the placement and location of DBS leads, as well as the extent of the corresponding VAT are pivotal determinants of various cognitive and emotional outcomes, no study to date has directly examined their relationship with FER. The present study therefore aims to elucidate whether and how changes in FER depend on the precise localisation of DBS leads and VAT within the STN.

Since DBS is known to influence impulsivity, which in turn may affect performance on FER tasks, we included an emotional impulsivity task in our analysis [19,20].

We found that facial emotion recognition depends on VAT location. We identified a lateralization of a positive effect of STN-DBS on recognition of positive emotions to the right hemisphere involving fibres connecting the STN to the prefrontal cortex. While we also identified altered valence processing where emotions were perceived as less negative, this effect was related to stimulation in the left dorsolateral STN, suggesting different mechanisms underlying DBS-induced alterations of the mere recognition of basic emotions and their perceived emotional affect.

## 2. Material and Methods

### 2.1. Ethical approval

This study was approved by the Ethics Board of the University Hospital Marburg (118/21) and adhered to the principles in the Declaration of Helsinki. Written informed consent was obtained from all participants prior to their inclusion.

### 2.2. Participants

A total of 54 participants were enrolled, divided into three groups: 22 healthy control subjects (HC), 16 PD patients without DBS and 16 PD patients undergoing bilateral DBS lead surgery. This report focuses on the DBS group, while demographic information and clinical outcomes of the other two groups are reported elsewhere [21]. The DBS group underwent bilateral STN-DBS lead surgery between March 2022 and July 2023 at the University Hospital Marburg following the established local protocols for pre-, peri-, and postoperative care [22,23]. All participants were evaluated twice: at a preoperative baseline visit and a post-operative follow-up three months [mean: 15.7 ± 3.72 weeks] after surgery to avoid influence of lesion effects.

### 2.3. Assessments and Questionnaires

Motor symptom severity was assessed using part III of the Movement Disorder Society Unified Parkinson’s Disease Rating Scale (MDS-UPDRS) [24]. Cognitive screening included the Montreal Cognitive Assessment (MoCA) [25] and the Frontal Assessment Battery (FAB) [26] for executive functions, both administered in the on-medication state. Signs of apathy, depressive or hypomanic symptoms, impulse control disorders, and PD-related quality of life were assessed using the Apathy Evaluation Scale (AES) [27], Beck’s Depression Inventory (BDI-II) [28], Hypomania Checklist 32 (HCL-32) [29], Questionnaire for impulsive-compulsive disorders in Parkinson’s Disease-Rating Scale (QUIP-RS) [30], and Parkinson’s Disease Questionnaire (PDQ-39) [31], respectively.

### 2.4. Task design

A range of tasks assessing emotion processing were administered after overnight withdrawal from dopamine replacement therapy (>12 hours), i.e., in the practically defined off-medication state. All tasks were programmed using the open-source software OpenSesame [32], with stimuli displaying basic facial emotions sourced from the Karolinska Directed Emotional Faces Database [33]. Non-emotional versions for each task were included to control for more general effects of changes in cognitive abilities post-surgery. Examples of the tasks described in this report are shown in Figure 1.

**Figure 1:**
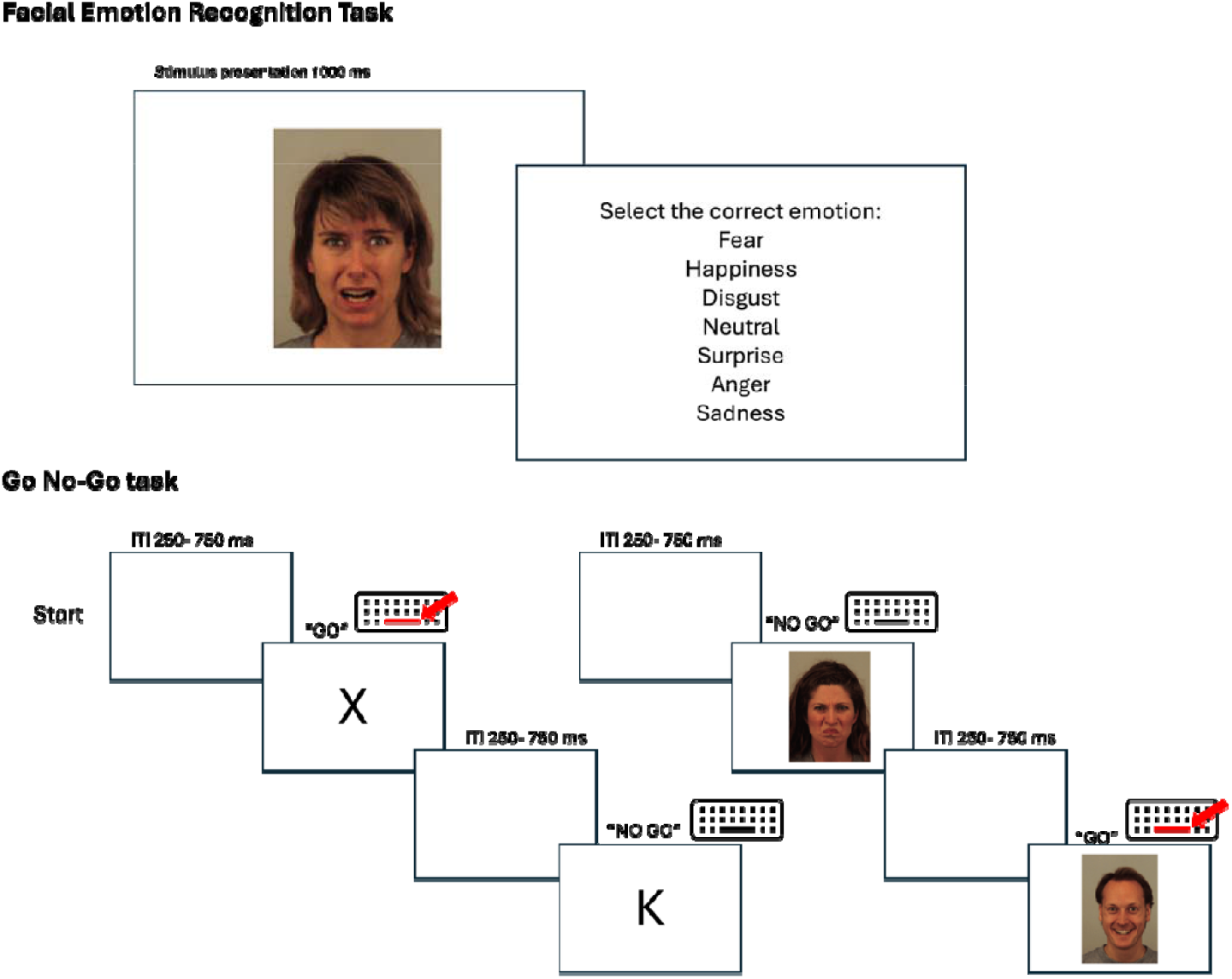
Task Design. Upper row: Facial Emotion Recognition task. Lower row: Tasks for testing impulsivity, with the left version testing the non-emotional component of the task by using the letters X and K as stimuli. Right: Emotional version of the task: Differentiation between happy and angry faces. The red line and arrow are visual representation of the button press required by the participant upon the previously introduced “Go” stimulus. No reaction was required in “No-Go” trials. In the original task, the version was presented in German instead of English which is used here for illustration purposes.

#### 2.4.1. Emotion recognition

In this task, participants identified one of seven basic emotions from static images of facial expressions: fear, disgust, happy, neutral, surprise, anger, or sadness. Each image was displayed for 1,000 ms, after which participants were asked to select the correct emotion from a list of options in a single-response, multiple-choice format. The identities of the stimuli varied across trials, with females and males represented equally. Two blocks of 77 pseudorandomised trials were presented with no time limit for responses, resulting in 22 trials per emotion. The emotional blocks were divided by a non-emotional version of the task in which participants were asked to identify the gender of displayed emotion (outcomes not reported here, please see [21]). Different versions were used for baseline and follow-up assessments. Outcomes of interest were overall accuracy pooled across all emotions and emotion-specific accuracy.

Given the limited number of trials per emotion and to avoid inflating alpha error due to multiple comparisons, we grouped emotions into positive and negative categories for analysis. However, recognising the ambiguous nature of “surprise”, we conducted a secondary analysis treating “happy” and “surprise” separately as described below. Building on prior studies suggesting that STN-DBS may differentially influence valence ratings of positive and negative emotions [34], we additionally examined whether VAT locations were associated with specific error patterns. To this end, we calculated the change in the proportion of trials in which a positive emotion was misclassified as negative and vice versa.

#### 2.4.2 Impulsivity

Emotional impulsivity was assessed using a Go No-Go task with a 70/30 contingency in which participants responded to static images of emotional faces. In the first block, they pressed the space key on a standard keyboard for happy faces and withheld for angry faces. This instruction was reversed in the second block. For the non-emotional version, either the letter “K” or “X” was used as Go and No-Go stimuli, respectively. Each block consisted of 50 trials, with participants given a maximum of 5000 ms per response. Outcome measures included accuracy (i.e., proportion of correct responses) and reaction time.

### 2.5. Visualization of Localisation of DBS leads and VAT

Image processing adhered to the pipeline proposed in the Lead-DBS software [35] as reported in several previous publications [8,22,36]. Briefly, aligned preoperative T1- and T2-weighted were co-registered to postoperative CT scans using two-stage linear interpolation method implemented in Advanced Normalization Tools. The co-registered images were then spatially warped to MNI space. Corrections for brain shift relied on a refined affine transformation between the pre- and post-operative images, focusing on subcortical areas of interest. The exact localisation of the DBS leads, and their orientation were determined using the PaCER algorithm [37]. Electrode locations and active contacts were visualized using Lead-Group [36,38].

For each patient’s DBS settings, the right and left VAT were estimated using a finite element method with an electric field threshold of 0.2 V/mm. This model distinguishes four compartments: grey matter, white matter, electrode contacts, and insulation. Grey matter structures, including STN subregion parcellation were defined using the DISTAL Minimal Atlas [39]. Patient-specific VAT were assessed for overlap with the dorsolateral motor and ventrolateral non-motor (associative and limbic area combined) subregions of the STN, expressed as proportions of the total VAT.

### 2.6. Whole-brain structural connectivity

Patient-specific VATs were used as seed regions within a normative connectome derived from diffusion-weighted MRI of 85 individuals with PD recruited for the Parkinson’s Progression Markers Initiative (PPMI) [39]. To investigate structural correlations of changes in FER, as proposed in the Lead-DBS pipeline and reported previously [35,40,41]. In brief, patient-specific VAT were projected onto the voxelized volume of the connectome in standard space in 1 mm isotropic resolution. Individual VAT was categorised based on their connectivity with each fibre of the connectome (connected vs. unconnected). A two-sample t-test was performed to compare FER changes between these two categories, with t-scores reflecting the explanatory power for each fibre for DBS-induced changes in FER. The top 20% of fibres with the highest t-scores were selected for further analysis.

### 2.7. Statistics and reproducibility

For all analyses, statistical inference was performed in R and statistical significance was asserted at α = 0.05. Performance on FER was evaluated using relative change scores [(post-pre)/pre]. Clinically significant improvement or decline at follow-up was defined as a change exceeding ± 1 SD from the group’s baseline mean. Spearman’s rank correlation was used to examine the relationship between VAT overlap and behavioural outcomes. Likewise, Spearman rank correlations were conducted to assess the relationship between fibre density in connected cortical voxels and improvements in FER. Here, a leave-one-patient-out cross-validation approach was employed.

## 3. Results

### 3.1. Participants’ clinical characteristics

In total, the DBS cohort comprised 16 right-handed patients (3 females) with a mean age of 60.3 ± 9.4 years and a mean disease duration of 7.5 ± 4.3 years. For additional clinical characteristics, we refer to Table 1.

**Table 1:** Clinical characteristics before and after DBS surgery.

| Time point | Baseline | 3-month follow-up |  |  |
| --- | --- | --- | --- | --- |
|  | Mean [SD] | Mean [SD] | p | T(15) |
| LEDD [mg] | 900.97 [313.36] | 625.19 [268.59] | 0.002 | 2.13 |
| UPDRS-III med on | 21.75 [9.58] | 12.08 [6.5]* | 0.01** | 2.12** |
| UPDRS-III med off | 37.56 [11.69] | 25.30 [11.55]* | 0.01** | 2.26** |
| MoCA | 27.88 [1.09] | 27.88 [1.41] | 1 | 2.13 |
| BDI-II | 7.69 [6.43] | 6.94 [6.93] | 0.55 | 2.13 |

\* STN-DBS was activated for both motor assessment at follow-up \*\*pre- and postoperative values were available in N= 13 for med-on and N= 10 for med-off

The mean time between the baseline study visit and DBS surgery was 2.2 ± 2.89 months. Subsequently, the follow-up visit was conducted 3.62 ± 0.86 months after surgery. The mean time between preoperative MRI and postoperative CT scans was 3.97 ± 1.00 months, and the time between CT scan and follow-up visit was 0.56 ± 1.36 months, respectively.

Lead visualisation confirmed that DBS leads were located in the target region, i.e., the dorsolateral aspect of the STN (Figure 2). As expected, DBS resulted in significant motor improvement (Table 1). No effects of VAT-STN overlap on post-operative changes in apathy, hypomania, depression, or quality of life were found in our cohort (Supplementary Table 1). Stimulation in the right motor aspect of the STN was associated with a slight improvement in performance in MoCA (ρ = 0.60, p = 0.014).

**Figure 2:**
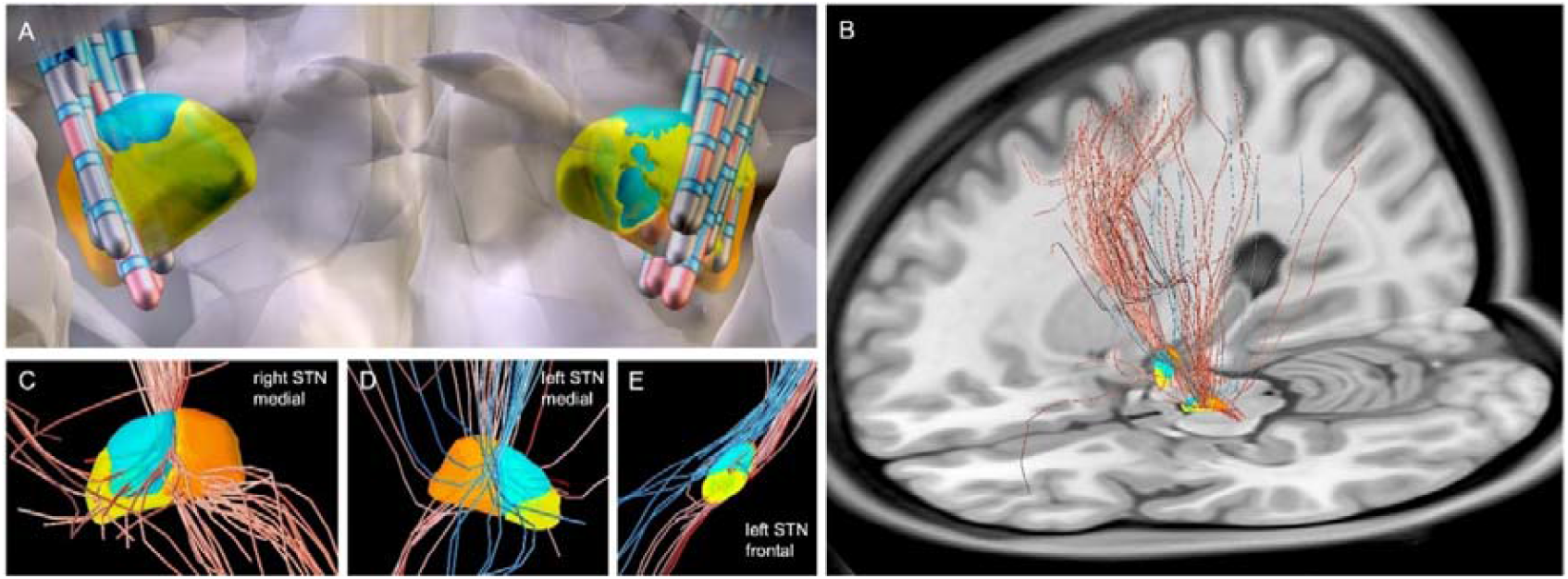
Visualisation of DBS leads and whole brain fibre tract analysis: A: Localisation of all DBS leads within the STN. Active contacts shown in red, inactive contacts in black. B: Tractography showing the results of the whole-brain fibre tract analysis with red fibres indicating a relationship with improved emotion recognition post-DBS and blue fibres indicating a relationship with decline in emotion recognition post-DBS, respectively. Only significant fibres are displayed. C-E: Close-ups of the right and left STN in medial view, respectively, and the left STN in frontal view. Positively correlated fibres (red tractography) appear to enter the non-motor region (blue associative part, blue limbic part) close to its dorsolateral intersection with the motor region (orange), whereas negatively correlated tracts bypass the left STN dorsolaterally (blue tractography).

### 3.2. Behavioural Tasks

#### 3.2.1. DBS-induced changes in Facial Emotion Recognition

On a group level, there was no evidence for a significant DBS-induced change in FER (F = 0.344, p = 0.557). Still, considerable variability between patients became evident. One participant showed overall improved FER (as defined by a change in accuracy > 1 SD of the group’s mean), while three declined. Similar pictures emerged when positive, negative, and neutral stimuli were analysed separately (Figure 3).

**Figure 3:**
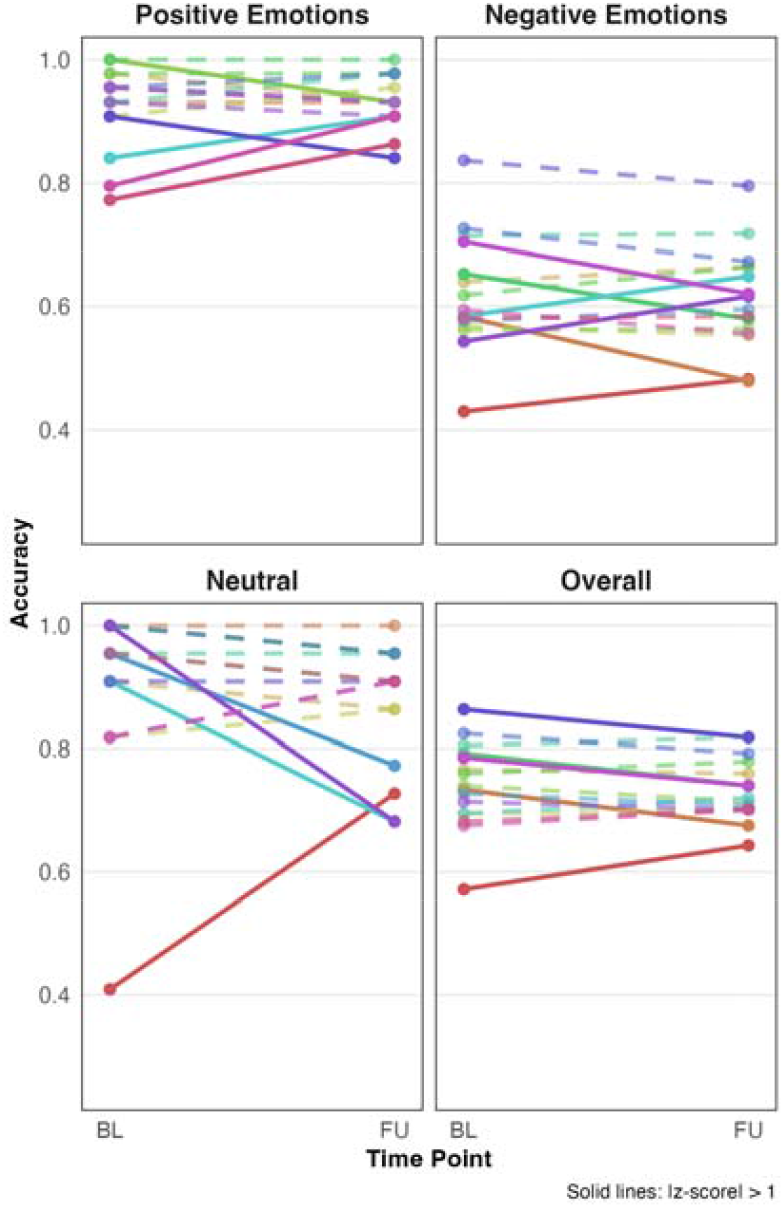
Individual changes in FER from baseline (BL) to follow-up (FU). Each coloured line represents the slope of one patient. Solid lines indicate that the individual’s absolute change exceeded one SD of the group’s mean, which was considered a clinically meaningful change. Dashed lines indicate smaller changes, respectively. Also note, the generally lower performance in recognizing negative versus positive emotions.

Stimulation of a larger portion of the ventrolateral non-motor part of the right STN correlated with improved emotion recognition after STN DBS (R = 0.54, p = 0.035) (Figure 2). When considering positive and negative emotions separately, it became apparent that this association was driven by improved recognition of positive emotions with stimulation in the non-motor STN (ρ = 0.54, p = 0.031), particularly in the right non-motor portion of the STN (ρ = 0.58, p =0.019), while no such correlation was found for negative emotions (ρ = 0.17, p = 0.53). Stimulation of the left motor STN, on the other hand, tended to be associated with a decrease in recognition of negative emotions, however, this did not reach statistical significance (ρ = -0.49, p = 0.056) (Figure 4).

**Figure 4:**
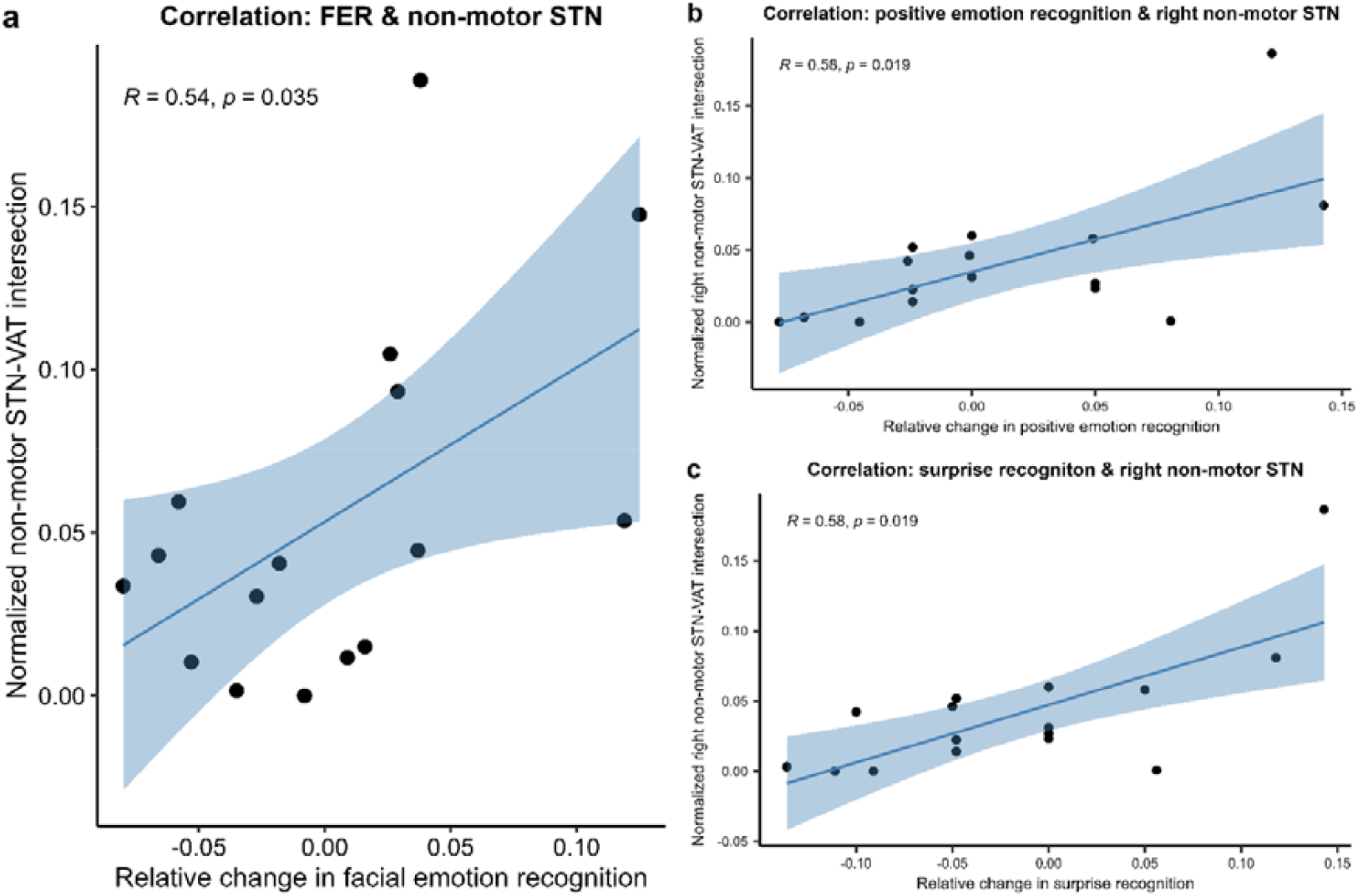
Spearman’s rank correlations between facial emotion recognition and DBS lead localisation within the STN. The blue shaded areas represent the 95% confidence interval.

**Figure 5:**
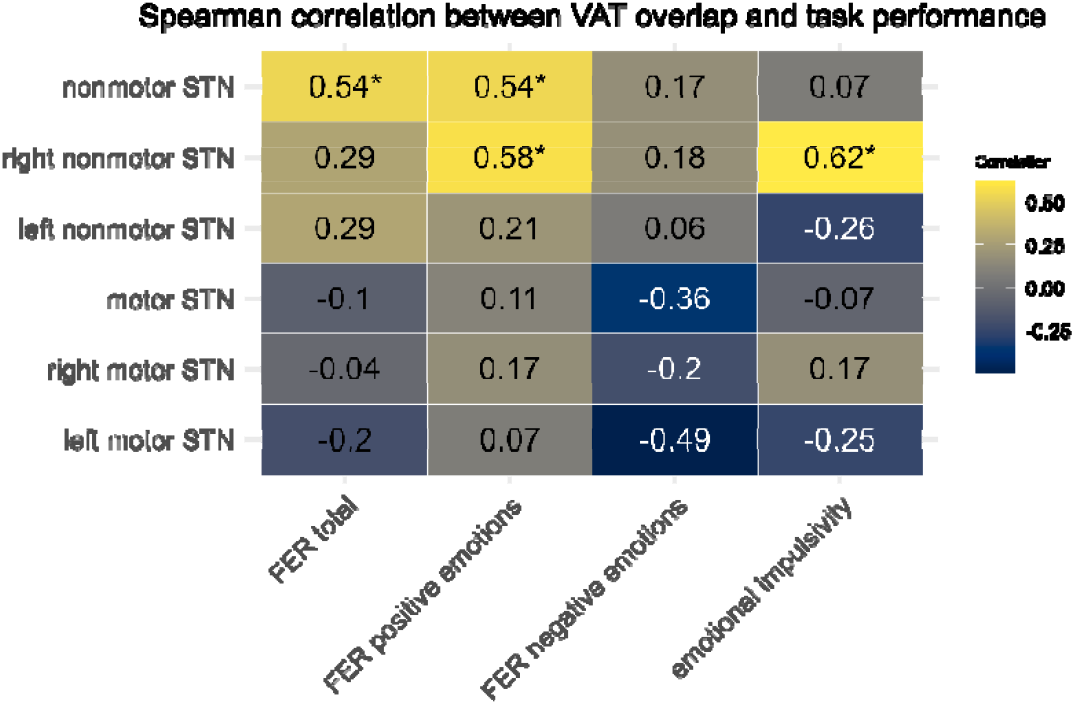
Spearman correlation between VAT intersection overlaps with proportion of the STN and task performance.

Given that surprise might not be considered an unambiguously positive emotion, we conducted a post-hoc analysis separating surprise and happiness recognition. Here, a particularly strong association of stimulation in the non-motor STN with improvement of surprise recognition (ρ= 0.63, p= 0.0085) became apparent, especially in the right hemisphere (ρ = 0.58, p = 0.019).

#### 3.2.2. Whole brain fibre tract analysis

The cross-validation leave-one-out approach was significant (ρ = -0.42, p = 0.04), implying that higher similarity of the individual set of discriminative fibres to the group fibre profile correlated with a larger DBS-induced effect on emotion recognition. Fibres associated with improvement in emotion recognition after DBS were right-dominant, crossing medial aspects of STN (**Figure 2 Panels B-C**), with most of them running through the dorsal aspects of the prefrontal cortex. Fibres associated with a decline in emotion recognition after DBS, on the other hand, tended to bypass the left STN dorsolaterally (**Figure 2 Panel D-E**). Analysis for positive and negative emotions separately, as well as for emotional impulsivity, did not yield significant results.

#### 3.2.3. DBS-associated Valence Changes in relation to VAT Location

There was a tendency to select more positive emotions when negative emotions were presented with a higher proportion of stimulation in the left motor STN, although this did not reach significance (R = 0.49, p = 0.052), but not the other way around (ρ = 0.14, p = 0.60) (**Supplementary Figure 1**). Given the relative similarity of the facial expressions of surprise and fear [42,43], we additionally checked for an association of VAT location with errors involving these two emotions. Indeed, there was a higher selection of surprise instead of afraid when a higher proportion of the VAT was in the motor STN, regardless of hemisphere (right: ρ = 0.54, p = 0.034; left: ρ = 0.67, p = 0.043). The reverse comparison showed no correlation, meaning that the participants did not confuse afraid with surprised facial expressions. These results indicate a potential bias towards perceiving emotions as less negative with stimulation in the dorsolateral part of the STN.

#### 3.2.4. DBS-induced Changes in Emotional Impulsivity

Higher proportions of VAT in the right non-motor STN were associated with reduced impulsivity as measured as ability to withhold responses to No-Go trials in the Go No-Go task. This effect was specific for emotional stimuli (ρ = 0.62, p = 0.01) as no such correlation was evident in the non-emotional task version (ρ = -0.055, p = 0.84).

## 4. Discussion

In this controlled prospective study, we show differential effects of STN-DBS on emotion recognition that are contingent upon the location of the stimulation volume within the STN. Specifically, we demonstrate that ventrolateral stimulation may improve processing of emotional stimuli, potentially underscoring the influence of cognitive and affective subcircuits within this central midbrain hub.

Recent literature indicates that STN-DBS surgery may provide non-motor symptom relief in PD patients [44–46]. The individual effects on distinct symptoms may hinge upon the location of the active electrodes [46,47]. The impact of STN-DBS on FER remains incompletely understood with previous behavioural studies yielding inconclusive results.

While ventral stimulation has been associated with improvements in mood [46] and anxiety [45,48] it is possible that DBS-related mood changes could influence our findings. However, it is important to emphasize that we did not observe significant changes in any of the behavioural scores, nor did we find a relationship between post-DBS changes in FER and changes in depressive symptoms or hypomania at an individual level [21]. Given the extensive connectivity of the STN with other brain regions, it is plausible that it interacts with structures involved in emotion processing, such as the ventral striatum, ventral pallidum, anterior cingulate cortex, and orbitofrontal cortex [3,49]. Metabolic studies indicate altered glucose metabolism following DBS in regions associated with FER, suggesting a modulation of networks between frontal and subcortical areas [12]. Building on these neuroanatomical and network-level considerations, our results point toward the importance of lead localisation and hemispheric lateralisation in shaping the emotional effects of DBS. Notably, we observed a lateralisation effect, emphasizing the dominance of the right hemisphere particularly its connections to the dorsolateral prefrontal cortex (DLPFC).

These findings align with two existing theories: (i) that the right hemisphere is dominant for the processing of emotions, and (ii) the processing of positive emotions is associated with the left prefrontal cortex, while negative emotions are linked to the right (valence-specific lateralisation hypothesis)[50]. Previous support for the right-hemisphere dominance model comes from studies demonstrating alpha desynchronisation selectively in the right STN in response to both positive and negative affect in PD [34]. Conversely, the valence-specific model is supported by evidence of distinct lateralisation patterns in the amygdala and prefrontal cortex [51–54]. While our findings support the right-hemisphere dominance hypothesis [55], they also suggest a more nuanced pattern, indicating that lateralisation may depend on the type of emotion and its connectivity with the prefrontal cortex.

Regarding different types of emotions, we found the strongest effect of VAT location on surprise recognition, a somewhat surprising finding, given that most prior studies reported the largest DBS-related effects on negative emotions [3,12]. However, the largest study to date (N=59), which found no overall change in FER one year after DBS, showed that surprise recognition exhibited the greatest individual variability, with both deterioration and improvement observed postoperatively [13]. However, surprise is not unequivocally positive and may signal unexpected or threatening events. Consistent with this, recent evidence suggests that the STN interrupts ongoing cognitive processes when confronted with unexpected stimuli [56] and regulates both the timing and quality of behavioural responses, thereby constraining impulsive actions [19,57]. In this context, surprising facial expressions might act as an alerting stimulus signalling the need to stop or adjust the current behaviour. A role of the STN for inhibition unwanted responses to emotional content is further supported by the fact that participants were less likely to engage in impulsive responses when confronted with emotional “No-Go” stimuli when a higher proportion of VAT was in the right non-motor STN, while we did not find no such correlations in the non-emotional condition.

The evaluation of images may reflect not only the recognition of the depicted emotions but also the subjective experience of these emotions. In addition to our findings indicating an association between ventral stimulation and improved emotion recognition, we observed that stimulation of the dorsolateral aspect of the STN was linked to a preference for selecting more positive emotions over neutral or negative ones, in particular for the relatively similar emotional expressions fear and surprise. This suggests a DBS-induced shift in valence processing which aligns with previous studies showing that STN-DBS may influence subjective emotional attribution depending on the stimulated area [34,58]. However, in contrast to our findings, a previous comparison of dorsal versus ventral stimulation within the STN in PD revealed that ventral stimulation selectively increased valence ratings of positive emotional images, while not affecting negative images [34,59]. Similarly, in individuals with obsessive-compulsive disorder (OCD), ventrolateral (non-motor) STN stimulation enhanced positive ratings and decreased negative ratings for both positive and negative emotional stimuli compared to the off-stimulation state [60]. While we also observe improvements in emotion recognition with stimulation in the non-motor, ventrolateral part of the STN, our results link dorsolateral stimulation sites to a “positivity bias” in valence ratings. Here, it is important to note that, in our study, only relatively small regions of the non-motor parts of the STN were stimulated, particularly in the most mediolateral part. These areas are known for their dense connections with the orbitofrontal cortex (OFC), and, consequently the limbic system, which plays a key role in FER. We observed that, for both positive and negative emotions, some fibres in the medial STN areas exhibited an inverse relationship compared to more lateral fibres, which are often associated with associative functions through the DLPFC. Therefore, based on our sample size and lead locations, we cannot exclude that stimulation of different areas within the non-motor aspect of the STN may yield different effects. A sample of individuals with OCD in which the medial part of the STN serves as the primary DBS target might be more suitable for addressing this question in the future.

An alternative explanation is that the improvement in FER observed with the stimuli used in our study may be linked more closely linked to modulation of the fronto-striatal associative pathway involving the DLPFC than to the limbic pathway. This observation suggests the possibility that improved FER in this context could result more from enhanced cognitive processing rather than from changes in emotional performance while solving those tasks. However, in our study, MoCA scores were consistently high at both baseline and follow-up, indicating stable cognitive functions among our patients.

## 5. Limitations

This study provides valuable insights into DBS-related changes in emotion recognition and their relationship to DBS lead localisation; however, several limitations warrant consideration. First, we used static rather than dynamic emotional stimuli. Because dynamic facial expressions are easier to recognise [3], this may have influenced accuracy. Nonetheless, as all participants were tested under identical conditions, the findings regarding electrode localisation remain valid. Second, we assessed emotion recognition three months after surgery. This timing may minimise the impact of lesion-related effects but should not capture long-term effects. Although a large study reported no significant changes in FER one-year post-DBS [13], longitudinal research is needed to clarify changes over time after lead surgery.

Given the relatively small sample size and exploratory design, we refrained from multiple comparison correction. Instead, we attempted to limit alpha error inflation by grouping emotional stimuli into negative and positive categories. Categorising surprise as a positive emotion may have been overly simplistic, since it can also evoke neutral or negative responses. Therefore, we conducted a secondary analysis to better distinguish it from happiness. Findings may also be influenced by the specific non-motor STN subregions stimulated. Certain areas may exert opposing effects on emotion recognition, highlighting the need for further investigation in more suitable samples such as patients with OCD.

The subdivision of the STN into functional subregions was accomplished through atlas-based methodology, which does not account for individual anatomical variation. Additionally, categorising the STN into two discrete functional zones oversimplifies the complex cortico-subthalamic circuitry. Although a dorsolateral-to-ventromedial organizational gradient exists, with motor functions predominantly dorsolateral and cognitive-limbic functions ventromedial, cortical projections overlap significantly throughout the STN. This convergence blurs boundaries between motor and non-motor territories. Nevertheless, this simplification has proven useful as a practical approximation of STN organisation in DBS research before. Finally, individual differences and external factors, including stimulus modality, task type, working memory, mood disorders and PD severity, may also contribute to variability in DBS outcomes [12,15]. Our small sample size precluded controlling for these potential confounders. Future studies with larger cohorts should address them to refine our understanding of DBS effects on affective processing.

## 6. Conclusion

In summary, our results indicate that changes in FER and emotional impulsivity under STN-DBS depend on stimulation site. Lateral stimulation of the non-motor STN was linked to improvements in these emotional capacities, underscoring the ventral STN’s role in emotion processing. This localisation-based perspective may help reconcile prior inconsistent findings on STN-DBS and provides a foundation for future research into non-motor side effects and their dependence on electrode placement.

## Data Availability

All data produced in the present study are available upon reasonable request to the authors.

## Acknowledgements

We would like to express our gratitude to all participants for their active involvement and valuable collaboration in this study. Special thanks are due to our study assistant Stefanie Spriewald for her assistance in recruiting participants.

## Author Contributions

CK: Investigation, Data curation, Formal analysis, Methodology, Visualization, Writing – original draft, **DB**: Investigation, Data curation, Writing – review & editing; **LT**: Resources, Supervision; **JW**: Conceptualization, Methodology, Visualization Software, Formal analysis, Writing – review & edition, Supervision; **DP**: Resources, Formal analysis, Writing – review & editing, Supervision.

