## Supplementary Material for "Variability of Changes in Emotion Recognition with Deep Brain Stimulation Depends on the Location of the Stimulation Volume in the Subthalamic Nucleus"

***Supplementary Table 1****: Spearman’s rank correlations for relative changes in FER, Impulsivity and questionnaires with VAT and overlap with STN non-motor and motor intersections.*

|  |  | **Overlap VAT**  **non-motor**  **STN**  **intersection** | | **Overlap VAT**  **right non-motor STN**  **intersection** | | **Overlap VAT**  **left non-motor**  **STN**  **intersection** | | **Overlap VAT**  **motor**  **STN**  **intersection** | | **Overlap VAT**  **right motor**  **STN**  **intersection** | | **Overlap VAT**  **left motor**  **STN**  **intersection** | |
| --- | --- | --- | --- | --- | --- | --- | --- | --- | --- | --- | --- | --- | --- |
|  |  | R | p | R | p | R | p | R | p | R | p | R | p |
| **Facial Emotion Recognition** | Total relative | **0.54** | **0.035** | 0.29 | 0.28 | 0.29 | 0.27 | -0.1 | 0.71 | -0.04 | 0.88 | -0.2 | 0.46 |
|  | Neg. emotions | 0.17 | 0.53 | 0.18 | 0.51 | 0.062 | 0.82 | -0.36 | 0.17 | -0.2 | 0.45 | ***-0.49*** | ***0.056*** |
|  | Pos. emotions | **0.54** | **0.031** | **0.58** | **0.019** | 0.21 | 0.44 | 0.11 | 0.69 | 0.17 | 0.54 | 0.074 | 0.79 |
|  | Afraid | 0.1 | 0.7 | 0.1 | 0.71 | 0.21 | 0.44 | -0.038 | 0.89 | 0.038 | 0.89 | -0.2 | 0.45 |
|  | Anger | -0.052 | 0.85 | -0.14 | 0.62 | -0.013 | 0.96 | 0.078 | 0.77 | 0.018 | 0.95 | 0.08 | 0.77 |
|  | Disgust | -0.11 | 0.68 | -0.19 | 0.49 | -0.28 | 0.29 | -0.007 | 0.98 | 0.035 | 0.9 | -0.096 | 0.72 |
|  | Happiness | 0.29 | 0.28 | 0.41 | 0.12 | 0.022 | 0.94 | 0.037 | 0.89 | 0.16 | 0.56 | -0.1 | 0.71 |
|  | Sad | 0.096 | 0.72 | 0.027 | 0.92 | 0.13 | 0.63 | -0.32 | 0.22 | -0.25 | 0.36 | -0.17 | 0.54 |
|  | Surprise | **0.63** | **0.0085** | **0.58** | **0.019** | 0.28 | 0.29 | 0.074 | 0.78 | 0.076 | 0.78 | 0.098 | 0.72 |
|  | Neutral | 0.31 | 0.24 | 0.043 | 0.87 | 0.32 | 0.22 | 0.17 | 0.54 | 0.058 | 0.83 | 0.095 | 0.73 |
| **Impulsivity** | No-Go accuracy (emotional) | 0.067 | 0.81 | **0.62** | **0.01** | -0.26 | 0.33 | -0.067 | 0.81 | 0.17 | 0.52 | -0.25 | 0.35 |
|  | No-Go accuracy (non emotional) | -0.13 | 0.64 | -0.06 | 0.84 | -0.2 | 0.47 | 0.11 | 0.69 | -0.019 | 0.95 | -0.018 | 0.95 |
|  | Time  (emotional) | 0.085 | 0.75 | 0.06 | 0.83 | -0.2 | 0.47 | -0.28 | 0.29 | -0.12 | 0.65 | -0.28 | 0.29 |
|  | Time  (non emotional) | -0.032 | 0.91 | 0.31 | 0.24 | -0.34 | 0.2 | 0.15 | 0.58 | 0.0621 | 0.82 | -0.021 | 0.94 |
| **Questionnaires** | MoCA | -0.18 | 0.49 | -0.22 | 0.42 | -0.087 | 0.75 | 0.44 | 0.088 | **0.6** | **0.014** | 0.28 | 0.27 |
|  | FAB | 0.21 | 0.44 | -0.039 | 0.89 | 0.17 | 0.53 | -0.17 | 0.53 | -0.27 | 0.31 | -0.19 | 0.47 |
|  | BDI.II* | -0.018 | 0.95 | 0.29 | 0.27 | -0.4 | 0.12 | -0.18 | 0.5 | -0.051 | 0.85 | -0.45 | 0.083 |
|  | AES | 0.1 | 0.7 | 0.078 | 0.77 | 0.27 | 0.31 | -0.043 | 0.88 | 0.023 | 0.93 | -0.1 | 0.71 |
|  | HCL_32* | -0.16 | 0.55 | 0.15 | 0.58 | -0.25 | 0.35 | -0.052 | 0.85 | 0.13 | 0.63 | -0.13 | 0.62 |
|  | PDQ_39* | -0.046 | 0.87 | -0.21 | 0.44 | -0.16 | 0.56 | -0.33 | 0.21 | -0.46 | 0.073 | -0.39 | 0.14 |
|  | QUIP.RS* | -0.15 | 0.59 | 0.02 | 0.95 | -0.32 | 0.22 | -0.05 | 0.85 | -0.032 | 0.91 | -0.17 | 0.53 |

***Supplementary Table 2: Valence of different emotions in correlation to overlap of different emotions (absolute****): Spearman’s rank correlation for selection of false emotion recognition in correlation to VAT overlap and STN intersections.*

| **chosen emotion** | **Correct**  **emotion** | **Overlap VAT**  **non-motor STN**  **intersection** | | **Overlap VAT**  **right non-motor STN**  **intersection** | | **Overlap VAT**  **left non-motor STN**  **intersection** | | **Overlap VAT**  **motor STN**  **intersection** | | **Overlap VAT**  **right motor STN**  **intersection** | | **Overlap VAT**  **left motor STN**  **intersection** | |
| --- | --- | --- | --- | --- | --- | --- | --- | --- | --- | --- | --- | --- | --- |
|  |  | R | p | R | p | R | p | R | p | R | p | R | p |
| Positive | Neg./ neu | -0.025 | 0.93 | 0.28 | 0.29 | -0.082 | 0.76 | 0.23 | 0.4 | 0.16 | 0.56 | 0.34 | 0.2 |
|  | Negative | -0.096 | 0.72 | 0.065 | 0.81 | -0.11 | 0.7 | 0.37 | 0.15 | 0.22 | 0.41 | ***0.49*** | ***0.052*** |
|  | afraid | -0.23 | 0.39 | 0.024 | 0.93 | -0.29 | 0.27 | **0.61** | **0.013** | ***0.49*** | ***0.053*** | **0.65** | **0.0064** |
|  | Neutral | 0.039 | 0.89 | **0.5** | **0.049** | -0.079 | 0.77 | -0.19 | 0.47 | -0.32 | 0.91 | -0.22 | 0.41 |
| Happy | Neg./ neu | -0.11 | 0.68 | -0.24 | 0.37 | 0.12 | 0.64 | -0.26 | 0.33 | -0.22 | 0.42 | -0.071 | 0.79 |
|  | Neutral | -0.28 | 0.29 | -0.056 | 0.84 | -0.36 | 0.17 | -0.035 | 0.9 | -0.13 | 0.62 | 0.065 | 0.81 |
|  | Negative | -0.22 | 0.42 | -0.34 | 0.19 | 0.069 | 0.8 | -0.33 | 0.21 | -0.34 | 0.2 | -0.14 | 0.6 |
|  | Afraid | -0.22 | 0.42 | -0.34 | 0.19 | 0.069 | 0.8 | -0.33 | 0.21 | -0.34 | 0.2 | -0.14 | 0.6 |
| Surprise | Neg./ neu | -0.024 | 0.93 | 0.35 | 0.18 | -0.16 | 0.56 | **0.53** | **0.036** | 0.45 | 0.082 | **0.53** | **0.034** |
|  | Neutral | -0.36 | 0.18 | 0.088 | 0.75 | -0.41 | 0.11 | 0.24 | 0.36 | 0.38 | 0.14 | 0.049 | 0.86 |
|  | Negative | -0.11 | 0.69 | 0.13 | 0.63 | -0.17 | 0.53 | 0.48 | 0.062 | 0.35 | 0.19 | **0.54** | **0.032** |
|  | Afraid | -0.13 | 0.64 | 0.11 | 0.69 | -0.25 | 0.36 | **0.66** | **0.0053** | **0.54** | **0.032** | **0.67** | **0.043** |
| Afraid | Surprise | -0.12 | 0.67 | -0.21 | 0.45 | 0.21 | 0.44 | -0.1 | 0.71 | -0.14 | 0.6 | -0.16 | 0.55 |
| Neutral | Negative | 0.42 | 0.1 | 0.14 | 0.6 | 0.23 | 0.4 | 0.04 | 0.88 | 0.016 | 0.95 | -0.069 | 0.8 |
|  | Positive | 0.33 | 0.21 | 0.066 | 0.81 | 0.2 | 0.46 | **-0.57** | **0.021** | -0.47 | 0.067 | **-0.7** | **0.0026** |
| Negative | Neutral | -0.38 | 0.14 | -0.16 | 0.55 | -0.3 | 0.26 | -0.15 | 0.58 | -0.084 | 0.76 | -0.048 | 0.86 |
|  | Positive | 0.25 | 0.35 | 0.08 | 0.76 | 0.34 | 0.2 | 0.08 | 0.76 | 0.03 | 0.92 | 0.14 | 0.60 |
